# Type I diabetes and incident dementia: a prospective study in the All of Us cohort

**DOI:** 10.1101/2025.07.11.25331402

**Authors:** Anna M Pederson, Peter Buto, Scott C Zimmerman, Mabeline Velez, Kendra D. Sims, Audrey R. Murchland, Jingxuan Wang, Alana T Brennan, M. Maria Glymour, Jennifer Weuve

## Abstract

**Importance:** Although diabetes mellitus (DM) is a well-established determinant of dementia risk, most studies have evaluated type 2 DM (T2DM) or any DM without considering type 1 DM (T1DM) separately. Questions remain about the influence of T1DM on risk of dementia.

**Objective:** To evaluate associations of T1DM and T2DM with incident dementia using linked electronic health records (EHRs).

**Design, Setting, Participants:** This cohort study used data from the All of Us (AoU) cohort, a convenience sample of US adults. Eligible participants were ≥ 50 years, completed baseline surveys, and had EHR information. Enrollment began in 2017, with data available through October 2023, including records prior to enrollment in AoU. Mean follow-up was 2.4 years.

**Exposures:** We developed and validated an algorithm to distinguish DM type using three reference measures: (1) self-report diabetes type; (2) C-peptide values; and (3) islet-specific autoantibodies (ISAs). Participants were classified as no DM, T1DM, or T2DM based on number of T1DM encounters.

**Main Outcomes and Measures:** Incident dementia was identified based on ICD-9, ICD-10, and SNOMED codes in participants’ EHRs.

**Results:** Among 283,965 participants (mean [SD] age 64.62 [8.96] years; 56.7% women); 60.3% identified as Non-Hispanic White; 13.3% as Hispanic/Latino; and 26.4% as Non-Hispanic Other. Optimal DM classification algorithm cutoas varied by reference standard: (1) self-reported diabetes: ≥ 1 T1DM EHR encounter (sensitivity: 0.59; specificity: 0.90); (2) C-peptide: ≥ 3 T1DM EHR encounters (sensitivity: 0.76; specificity: 0.79); and (3) ISAs: ≥ 4 T1DM EHR encounters (sensitivity: 0.48; specificity: 0.74). Using at least one T1DM encounter cutoa, 5,444 participants were classified with T1DM. Compared with those without DM, participants with T1DM had higher incidence of dementia (sociodemographic-adjusted HR = 2.79; 95% CI: 2.26-3.45); those with T2DM also had elevated risk (sociodemographic-adjusted HR = 2.09; 95% CI: 1.88-2.33). Results were similar across gender and race and ethnicity stratified groups.

**Conclusion and Relevance:** In this cohort, participants with diabetes had a higher dementia risk than did those without DM, with the highest risk among those with T1DM. These findings highlight the need to better understand mechanisms linking T1DM and dementia in aging populations.

## Introduction

Diabetes mellitus (DM), a leading cause of morbidity and mortality, has well-documented links to dementia. ^1–3^ Although type 2 diabetes (T2DM) accounts for approximately 95% of diabetes cases, the far less prevalent type 1 diabetes (T1DM) has historically led to reduced life expectancy.^4–7^ Because many studies fail to differentiate between the far less prevalent type 1 diabetes (T1DM) and T2DM or focus only on individuals with T2DM, the risk of dementia associated with T1DM remains unclear.^8–12^

Advances in T1DM treatment have extended many individuals’ lives such that they face aging-related diseases, including dementia.^13^ Existing studies of the association of T1DM with dementia have notable limitations such as limited diversity, retrospective study designs, small numbers of individuals with T1DM, or focus on other exposures within T1DM populations without directly assessing T1DM itself as a risk factor.^14–22^ Given the increasing number of aging individuals with T1DM, it is crucial to understand the relation of T1DM to dementia risk.

T1DM is rare, and electronic health records (EHRs) offer a critical opportunity to research T1DM and dementia in large samples. However, methods for accurately identifying individuals with T1DM using EHRs are not well developed. In EHR-based cohorts, distinguishing between T1DM and T2DM may be challenging due to overlaps in diagnostic tests and co-occurring T1DM and T2DM.

Using subsamples of a large EHR cohort, we validated an algorithm using three reference standard measures for T1DM to identify T1DM cases from EHR encounters. We then examined the association between DM type and dementia incidence, overall and by race, ethnicity and gender.

## Methods

### Study Sample

All of Us (AoU) is an ongoing longitudinal cohort of individuals ages 18 and older recruited as a convenience sample from across the United States.^23^ Recruitment began in 2017 through health care provider organizations and volunteer-based enrollment (eTable 5 in the Supplement).^24^ At enrollment, participants complete baseline surveys on sociodemographic characteristics and health needs. All AoU participants are asked for consent to link to their EHRs; however, providing this consent is optional.^23,24^ Follow-up information for our study was available through October 10, 2023. We followed the Strengthening of the Reporting of Observational Studies in Epidemiology (STROBE) reporting guideline for cohort studies.^25^

### Algorithm for Classifying Diabetes Type

In the absence of uniformly available diagnostic data, we developed a measure to distinguish individuals with T1DM from those with T2DM based on clinical encounter data. EHR encounters were coded based on ICD-9, ICD-10, and corresponding Systematized Nomenclature of Medicine (SNOMED) codes (eTable 8 in the Supplement). To evaluate the validity of the EHR-based classification, we leveraged laboratory-measured and self-reported diabetes type (eMethods and eTable 6 in the Supplement).

### Reference standards

For identifying persons with T1DM, three measures, each available for a subset of the full sample, defined our reference standards; (1) self-reported T1DM or T2DM (whether a participant indicated they had T1DM and/or T2DM on the personal and family history survey); (2) C-peptide concentration (T1DM: ≤ 0.20 nmol/L vs T2DM: > 0.20 nmol/L); and (3) islet-specific autoantibodies (ISA) positivity (T1DM: antibody > 0). Self-reported information is commonly used in large cohort studies and can oaer additional context when clinical diagnostic information data are unavailable. C-peptide and ISAs are widely used diagnostic data for identifying T1DM. C-peptide reflects pancreatic beta cell function; ISAs indicate autoimmune destruction of beta cells, a hallmark of T1DM.^26,27^

### Study sample for developing the classification algorithm

To develop the classification algorithm, we extracted data on all AoU participants, ages 18+ at enrollment, with an EHR record of diabetes (either T1DM or T2DM) who also had at least one reference standard measured: self-reported diabetes (n=19,718), C-peptide measurement (n=973), or ISA measurement (n=603) (eTable 1 in the Supplement).

### Statistical analyses for developing the classification algorithm

For each participant with DM EHR encounters, we separately counted the number of T1DM and T2DM EHR encounters. Among participants with DM, we determined the number of T1DM encounters that maximized the sensitivity and specificity of classifying participants with T1DM (vs T2DM) against each of the three reference standards, as follows (eFigure 1 in the Supplement): we set a threshold of T1DM EHR encounters above which participants were classified as having T1DM and as having T2DM otherwise (irrespective of the number of EHR encounters for T2DM). We calculated sensitivity and specificity over a range of thresholds from 0 to 50 to systematically evaluate the algorithm’s performance and choose an optimal threshold. For each threshold, we calculated: sensitivity and specificity for T1DM. We additionally defined the optimal threshold for each reference measure as the threshold that maximized Youden’s J-index, calculated using the cutpointr package in R.^28–30^

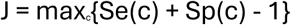

**Figure 1.**
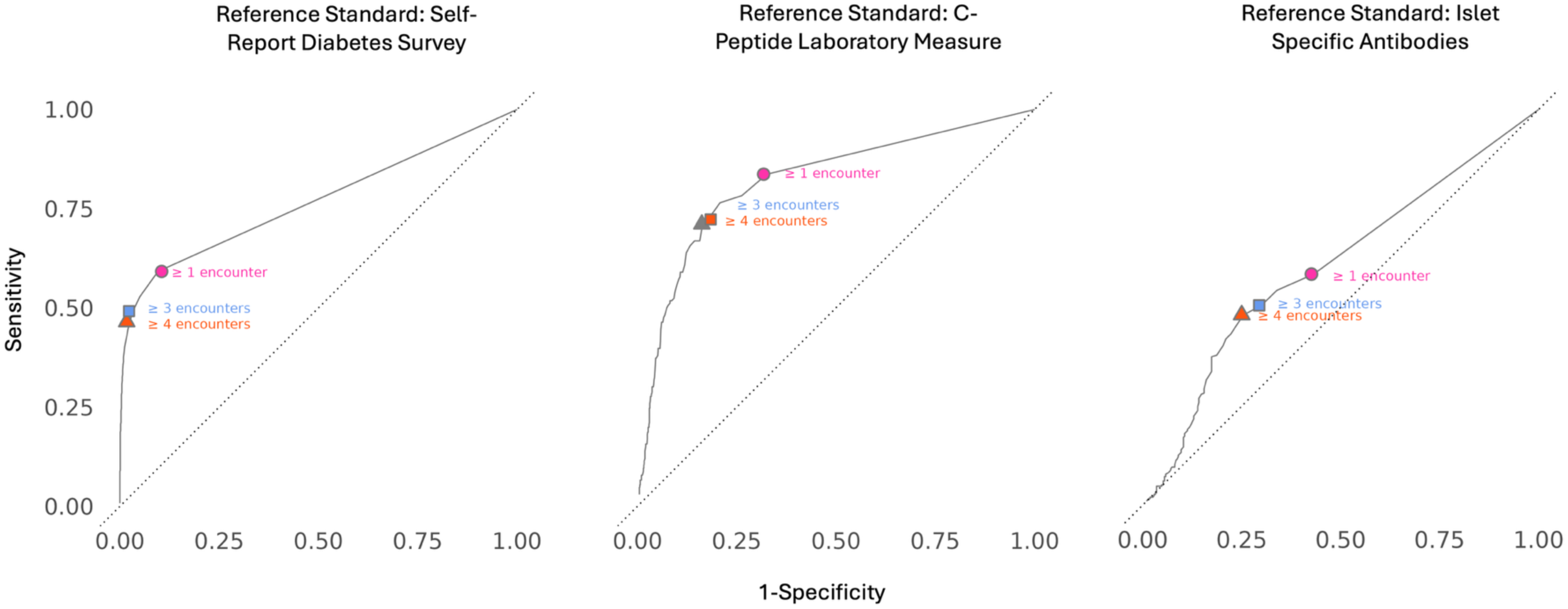
Sensitivity and specificity of type 1 diabetes classification (vs type 2 diabetes) of 200 thresholds of EHR encounters for diabetes, among All of Us participants with diabetes Each plot corresponds to the accuracy of given EHR encounter thresholds in predicting type 1 diabetes as defined by (1) self-report, (2) C-peptide levels, and (3) islet-specific antibodies (ISA). The sensitivity and specificity of ≥1 EHR encounter were: (1) self-reported diabetes: sensitivity = 0.59; specificity = 0.90; (2) C-Peptide levels: sensitivity = 0.83; specificity = 0.67; and (3) ISA: sensitivity = 0.58; specificity = 0.56. The sensitivity and specificity of ≥3 EHR encounters were: (1) self-reported diabetes: sensitivity = 0.49; specificity = 0.96; (2) C-Peptide levels: sensitivity = 0.76; specificity = 0.79; and (3) ISA: sensitivity = 0.50; specificity = 0.70. The sensitivity and specificity of ≥4 EHR encounters were: (1) self-reported diabetes: sensitivity = 0.46; specificity = 0.97; (2) C-peptide levels: sensitivity = 0.72; specificity = 0.82; and (3) ISA: sensitivity = 0.48; specificity = 0.74.

The optimal cutoff, J, is the number of T1DM EHR encounters (c) that maximizes the sum of sensitivity (Se) and specificity (Sp).

### Estimation of the e:ect of T1DM on dementia incidence

#### Study sample

Our analyses included AoU participants who were 50 years or older at enrollment and who had EHR linkage. We excluded 2,126 individuals with a diagnosis of dementia in their EHR prior to completion of this survey.

#### Assessment of T1DM and T2DM

Participants with no DM EHR encounter were classified as not having DM. Using our diabetes classification algorithm, we classified those with any diabetes encounter into diabetes types T1DM or T2DM. Individuals were classified as T1DM if they had at least one encounter for T1DM (the threshold we deemed most optimal based on our algorithm validation [see Results Section *Validation of the Algorithm for Classifying Diabetes Type* and eTable 2 in the Supplement]); those with no T1DM encounter were classified as having T2DM. Participants categorized as both T1DM and T2DM were grouped with the T1DM category due to small sample sizes.

#### Assessment of All-Cause Dementia

All-cause dementia included Alzheimer’s dementia (AD), vascular dementia, and dementia of unknown etiology (eTable 7 in the Supplement). We did not include diagnosis of frontotemporal, Lewy Body, or alcohol-related dementias, as these conditions may preclude the diagnosis of other forms of dementia. We defined time to incident all-cause dementia as time from baseline to the first appearance in a participant’s EHR of a corresponding ICD-9, ICD-10, or SNOMED code.

Individuals were censored upon dementia diagnosis, death, or the administrative censoring date defined by the last available EHR encounter date in the sample (October 1, 2023).

#### Covariates

We estimated eaects of each type of DM on dementia incidence adjusting for two sets of covariates representing potential sources of confounding. All covariate data were self-reported at baseline.

The first covariate set included sociodemographic factors: age at baseline, gender (man/woman), racial and ethnic identity (Non-Hispanic White, Non-Hispanic Black, Non-Hispanic Asian, Hispanic/Latino, or Non-Hispanic Other), educational attainment (self-reported highest level of school completed: less than high school, high school, some college, or college and above), and household income (adjusted by dividing by the square root of household size). The second set additionally included behaviors at midlife that, depending on their timing relative to DM onset, could be sources of confounding or responses to a DM diagnosis. These included: smoking history (never smoked or ever smoking at least 100 cigarettes in your life), and alcohol use (never, past, low-use, and high-use, based on the National Institute on Alcohol Abuse and Alcoholism (NIAAA) risk categories that account for frequency, quantity, and gender).

#### Statistical Analyses

We used Cox proportional-hazards regression models, using time from baseline to dementia as the time scale, to estimate hazard ratios for the association of DM type, versus no DM, with all-cause dementia. Following baseline, participants with no recorded EHR encounter for 448 days (the 99th percentile of the distribution of days between encounters) were assumed to no longer be receiving care from a clinic with linked EHR data and were censored at 448 days after the date of their last encounter. We fit: (1) a model adjusted for sociodemographic factors (set 1); and (2) a model adjusted for both sociodemographic- and mid-life factors (set 2). We also conducted analyses stratified by race and ethnicity and gender.

To protect participant privacy, we suppressed results based on fewer than 20 dementia events. Given small participant numbers in some racial and ethnic groups, we combined the Asian and Black subgroups with the Other race and ethnicity stratum.

To quantify the proportion of the dementia incidence among those with T1DM that can be attributed to T1DM, we calculated the attributable fraction among those with T1DM, as HR-1/HR.^31^ To quantify the contribution of T1DM to the incidence of all-cause dementia in our analytical sample, we estimated the following population attributable fraction (PAF), using Miettinen’s formula:^32,33^

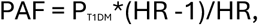

where P_T1DM_ is the prevalence of T1DM among individuals who developed dementia in our analytical sample, and HR is the estimated T1DM-dementia hazard ratio from the model adjusted for sociodemographic factors (set 1).

#### Sensitivity analyses

In sensitivity analyses, we defined T1DM as having at least three T1DM encounters (instead of at least 1). This definition had higher specificity, according to the reference standards used to validate our classification algorithm. For individuals with fewer than three T1DM encounters, classification was based on the proportion of their total DM encounters that were for T1DM. If this proportion was greater than or equal to 0.5, they were classified as having T1DM; if this proportion was less than 0.5, they were classified as having T2DM.

## Results

### Validation of the Algorithm for Classifying Diabetes Type

Using Youden’s J-index, the optimal threshold of EHR encounters for classifying T1DM versus T2DM varied across the three reference standard measures (Figure 1; eTable 2 in the Supplement): ≥ 1 encounter for self-reported diabetes type (sensitivity: 0.59; specificity: 0.90); ≥ 3 encounters for C-peptides (sensitivity: 0.76; specificity: 0.79); and ≥ 4 encounters for ISAs (sensitivity: 0.48; specificity: 0.74).

### Associations of diabetes type with incident dementia

Among the 283,965 participants in the analyses of DM and dementia, 5,444 (1.91%) were classified as having T1DM at baseline, based on having at least one T1DM EHR encounter, and 51,536 (18.14%) were classified as having T2DM. Over a mean of 2.36 years of follow-up, 2,361 participants developed dementia. The mean baseline age was 64.6 years (SD = 8.9), 56.7% were female, and 47.1% had at least a college degree (Table 1).

**Table 1.** Characteristics of the analytic sample, by diabetes status.

|  | All (n=283965) | Type 1 DM<br>(n=5444) | Type 2 DM<br>(n=51536) | No Diabetes<br>(n=226985) |
| --- | --- | --- | --- | --- |
| **Age (years), mean (SD)** | 64.62 (8.96) | 65.11 (8.72) | 64.95 (8.85) | 64.53 (8.99) |
| **Female, n (%)** | 160934 (56.7%) | 2742 (50.4%) | 27791 (53.9%) | 130401 (57.4%) |
| **Race and ethnicity, n (%)** |  |  |  |  |
| White | 171156 (60.3%) | 2598 (47.7%) | 25174 (48.8%) | 143384 (63.2%) |
| Hispanic/Latino | 37816 (13.3%) | 1030 (18.9%) | 9523 (18.5%) | 27263 (12%) |
| Other | 74993 (26.4%) | 1816 (33.4%) | 16839 (32.7%) | 56338 (24.8%) |
| **Educational attainment, n (%)** |  |  |  |  |
| Advanced Degree | 69624 (24.5%) | 826 (15.2%) | 7979 (15.5%) | 60819 (26.8%) |
| College Graduate | 64152 (22.6%) | 1059 (19.5%) | 9438 (18.3%) | 53655 (23.6%) |
| Some College | 72176 (25.4%) | 1590 (29.2%) | 15137 (29.4%) | 55449 (24.4%) |
| High School or GED | 46658 (16.4%) | 1074 (19.7%) | 10909 (21.2%) | 34675 (15.3%) |
| Less than High School | 24068 (8.5%) | 739 (13.6%) | 6662 (12.9%) | 16667 (7.3%) |
| **BMI (kg/m<sup>2</sup>), mean (SD)** | 29.83 (7.06) | 31.99 (7.47) | 32.87 (7.53) | 28.83 (6.6) |
| **Income (divided by the square root of household size), mean (SD)** | 57255.69<br>(50010.69) | 40399.9<br>(41427.05) | 41986.25<br>(41733.39) | 60815.82<br>(51081.54) |
| **Ever smoked (at least 100 Cigarettes in lifetime), n (%)** | 122955 (43.3%) | 2474 (45.4%) | 24086 (46.7%) | 96395 (42.5%) |
| **Alcohol Use, n (%)** |  |  |  |  |
| Never | 25355 (8.9%) | 746 (13.7%) | 6431 (12.5%) | 18178 (8%) |
| Past | 54175 (19.1%) | 1638 (30.1%) | 13446 (26.1%) | 39091 (17.2%) |
| Low | 70417 (24.8%) | 817 (15%) | 8700 (16.9%) | 60900 (26.8%) |
| High | 49050 (17.3%) | 571 (10.5%) | 6121 (11.9%) | 42358 (18.7%) |

T1DM (versus no DM) was associated with much higher incidence of dementia, whether adjusted for sociodemographic factors dementia (HR = 2.79; 95% CI: 2.26-3.45) or also for mid-life factors (HR = 2.37; 95% CI: 1.78-3.07) (Table 2). T2DM was also associated with higher dementia risk, compared with not having DM, (adjusted for sociodemographic factors, HR = 2.09, 95% CI: 1.88-2.33; additionally adjusted for mid-life factors, HR = 2.02, 95% CI: 1.78-2.29).

**Table 2.**
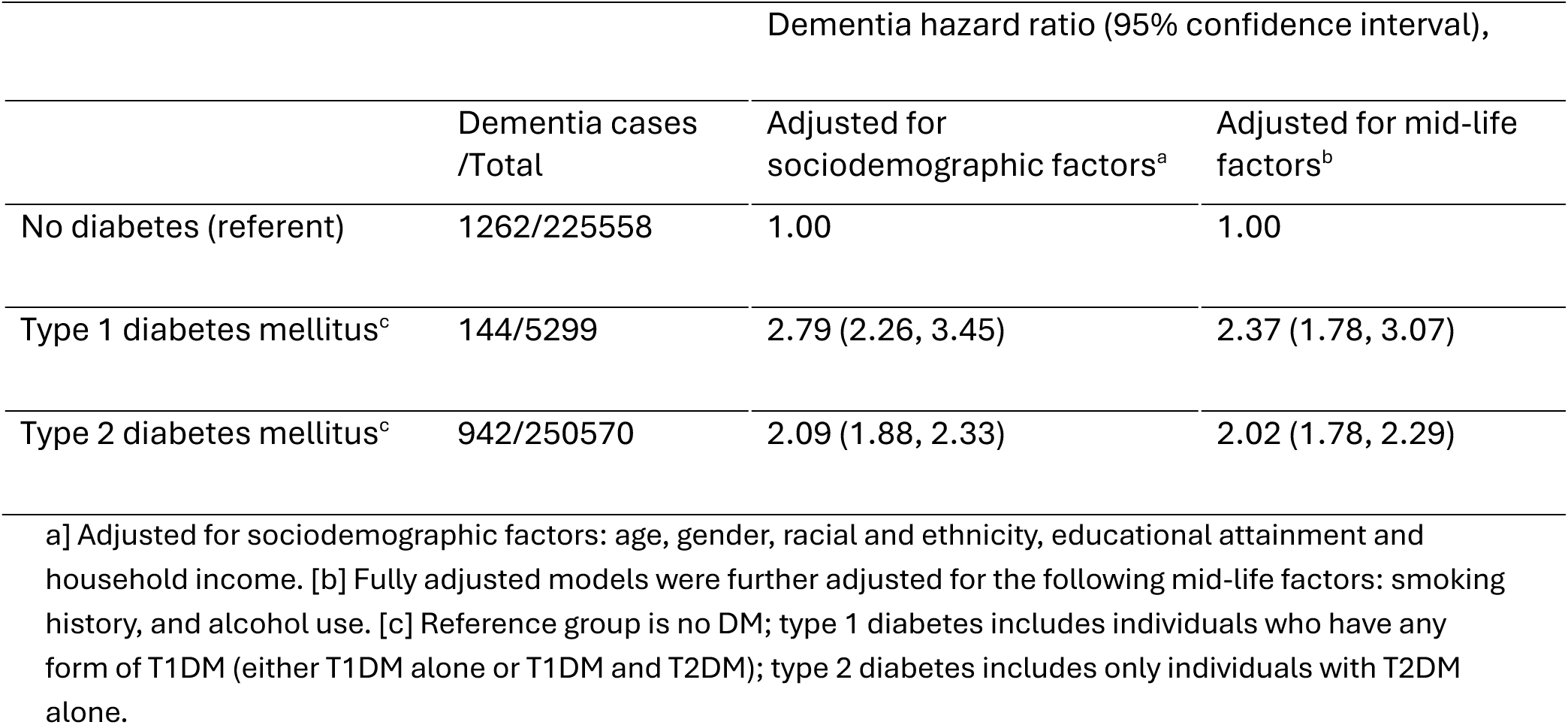
Dementia hazard ratio associated with each type of diabetes mellitus (reference: no diabetes mellitus), all participants.

### Gender-stratified associations

T1DM was associated with higher hazard of dementia among females (set 1 HR = 3.04; 95% CI: 2.28-4.05; Table 3) and males (set 1 HR = 2.59; 95% CI: 1.90-3.54). Among both females and males, T2DM was associated with approximately double the dementia hazard rate, compared with not having DM.

**Table 3.**
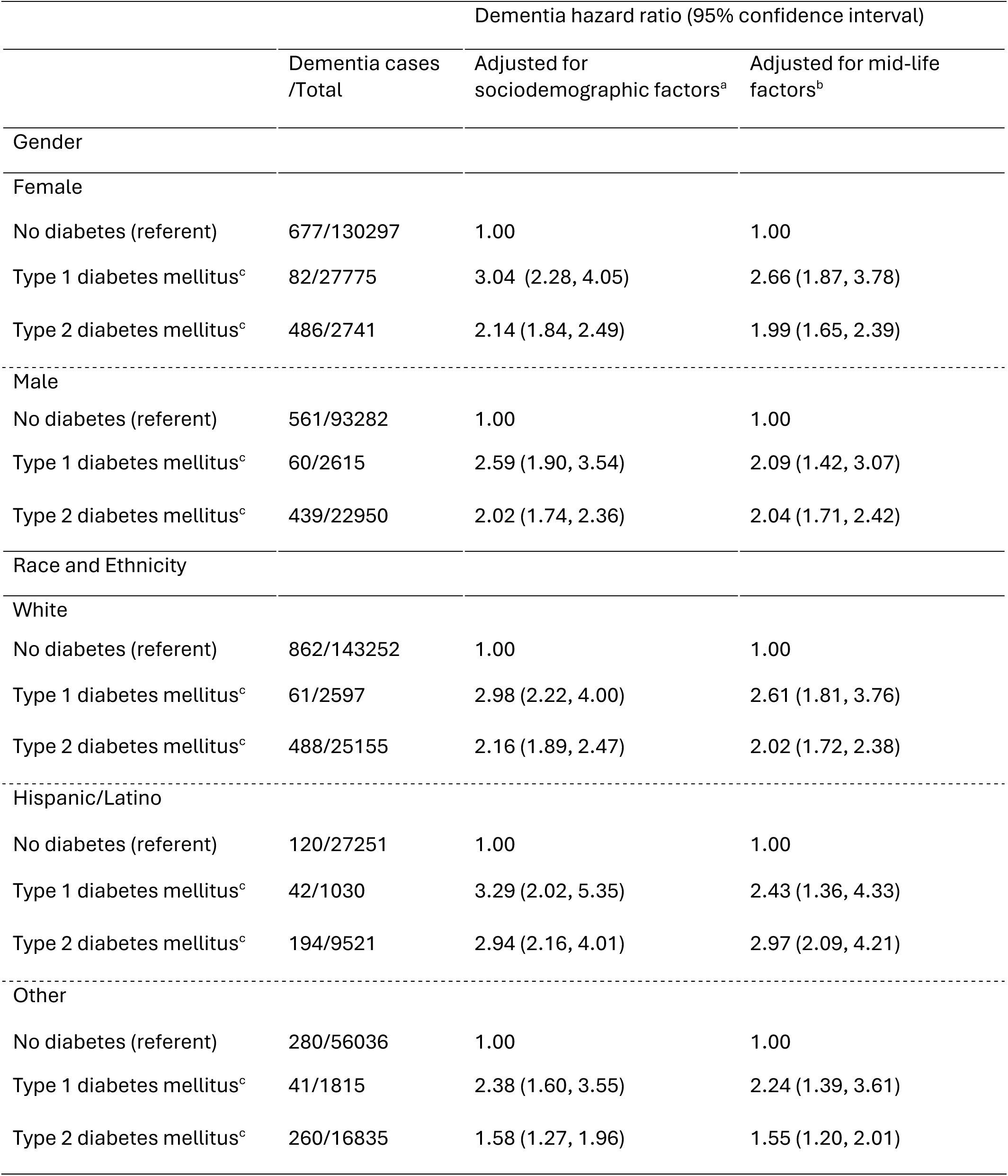
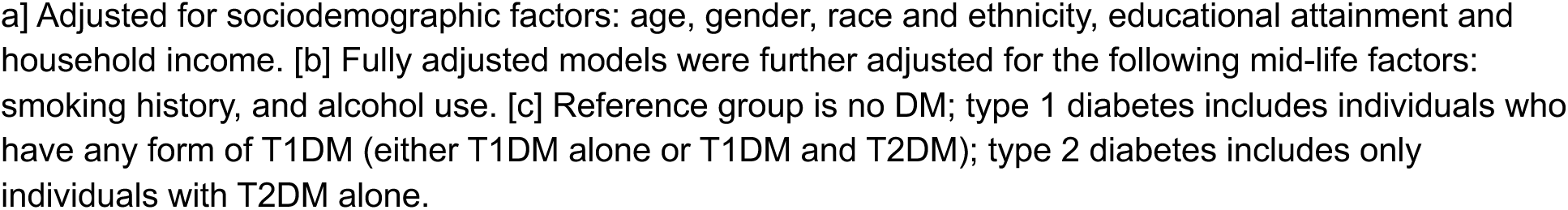
Dementia hazard ratio associated with each type of diabetes mellitus (reference: no diabetes mellitus), within strata of gender, and race and ethnicity.

|  |  | Dementia hazard ratio (95% confidence interval) |  |
| --- | --- | --- | --- |
|  | Dementia cases /Total | Adjusted for sociodemographic factors<sup>a</sup> | Adjusted for mid-life factors<sup>b</sup> |
| Gender |  |  |  |
| Female |  |  |  |
| No diabetes (referent) | 677/130297 | 1.00 | 1.00 |
| Type 1 diabetes mellitus<sup>c</sup> | 82/27775 | 3.04 (2.28, 4.05) | 2.66 (1.87, 3.78) |
| Type 2 diabetes mellitus<sup>c</sup> | 486/2741 | 2.14 (1.84, 2.49) | 1.99 (1.65, 2.39) |
| Male |  |  |  |
| No diabetes (referent) | 561/93282 | 1.00 | 1.00 |
| Type 1 diabetes mellitus<sup>c</sup> | 60/2615 | 2.59 (1.90, 3.54) | 2.09 (1.42, 3.07) |
| Type 2 diabetes mellitus<sup>c</sup> | 439/22950 | 2.02 (1.74, 2.36) | 2.04 (1.71, 2.42) |
| Race and Ethnicity |  |  |  |
| White |  |  |  |
| No diabetes (referent) | 862/143252 | 1.00 | 1.00 |
| Type 1 diabetes mellitus<sup>c</sup> | 61/2597 | 2.98 (2.22, 4.00) | 2.61 (1.81, 3.76) |
| Type 2 diabetes mellitus<sup>c</sup> | 488/25155 | 2.16 (1.89, 2.47) | 2.02 (1.72, 2.38) |
| Hispanic/Latino |  |  |  |
| No diabetes (referent) | 120/27251 | 1.00 | 1.00 |
| Type 1 diabetes mellitus<sup>c</sup> | 42/1030 | 3.29 (2.02, 5.35) | 2.43 (1.36, 4.33) |
| Type 2 diabetes mellitus<sup>c</sup> | 194/9521 | 2.94 (2.16, 4.01) | 2.97 (2.09, 4.21) |
| Other |  |  |  |
| No diabetes (referent) | 280/56036 | 1.00 | 1.00 |
| Type 1 diabetes mellitus<sup>c</sup> | 41/1815 | 2.38 (1.60, 3.55) | 2.24 (1.39, 3.61) |
| Type 2 diabetes mellitus<sup>c</sup> | 260/16835 | 1.58 (1.27, 1.96) | 1.55 (1.20, 2.01) |

a] Adjusted for sociodemographic factors: age, gender, race and ethnicity, educational attainment and household income. [b] Fully adjusted models were further adjusted for the following mid-life factors: smoking history, and alcohol use. [c] Reference group is no DM; type 1 diabetes includes individuals who have any form of T1DM (either T1DM alone or T1DM and T2DM); type 2 diabetes includes only individuals with T2DM alone.

### Race and ethnicity-stratified associations

T1DM was associated with elevated dementia incidence in all three race and ethnicity groups (HR_Hispanic/Latino_= 3.29; 95% CI: 2.02-5.35; HR_White_= 2.98; 95% CI: 2.22-4.00, HR_Other_ = 2.38; 95% CI: 1.60-3.55; Table 3). These associations changed little after adjustment for mid-life factors (set 2).

In all racial and ethnic groups, T2DM was associated with higher dementia risk (HR_Hispanic/Latino_= 2.94; 95% CI: 2.16-4.01; HR_White_= 2.16; 95% CI: 1.89-2.47, HR_Other_ = 1.58; 95% CI: 1.27-1.96; Table 3).

These associations remained after adjustment for mid-life factors (Table 3).

### Attributable Fractions and Population Attributable Fractions

With a T1DM prevalence of 6.05% among individuals who developed dementia, we estimated that 3.90% of dementia cases among AoU participants and 64.5% of dementia cases among people with T1DM could be attributed to T1DM (eTable 4 in the Supplement).

### Sensitivity Analyses

Estimated associations of DM with dementia incidence were similar when we classified T1DM based on a stricter criterion–requiring more than 3 T1DM EHR encounters (eTable 3 in the Supplement). Under this stricter definition, 2,787 participants were classified as having T1DM. Compared with individuals without DM, dementia incidence was higher among those with T1DM (HR = 2.66; 95% CI: 1.98-3.55) and among those with T2DM (HR = 1.93; 95% CI: 1.73-2.15), adjusting for sociodemographic factors. Gender-stratified results were also similar (female T1DM HR adjusted for sociodemographic factors = 2.74; 95% CI: 1.84-4.08; male T1DM HR = 2.54; 95% CI: 1.66-3.90; Table 3). Due to small sample sizes, we did not stratify by race and ethnicity.

## Discussion

We developed an algorithm for identifying the optimal diabetes encounters to classify AoU participants with T1DM based on EHR encounters. Both T1DM and T2DM were associated with more than twice the risk of dementia compared with not having DM with moderately greater elevation in risk associated with T1DM. Estimates were similar across gender and race and ethnicity.

A previous algorithm developed in the AoU cohort used EHR data, including laboratory results and prescription medications, as well as polygenic scores and self-reported diabetes status.^34^ In this algorithm, insulin use was a criterion for identifying individuals with T1DM, yet many individuals with T2DM use insulin in advanced stages of the disease.^35^ Our approach builds upon this work by validating DM type against three reference standards, including self-report and two clinical biomarkers that provide a biologically grounded framework for distinguishing T1DM from T2DM. This distinction may be particularly important when evaluating outcomes, like dementia, that may have etiologic pathways specific to DM type.

Our findings advance the existing evidence that DM is related to higher risk of dementia.^8–12^ In spite of the vast evidence amassed, there remains a need to evaluate dementia risk by DM type. For example, the 2024 Lancet Commission report on dementia prevention prioritizes diabetes as a modifiable risk factor for dementia, but this is primarily based on meta-analyses of studies evaluating the effects of T2DM.^36^ Thus, it remains important to understand the impact of T1DM on dementia risk, especially as life expectancy for individuals with T1DM lengthens. Our study offers valuable evidence by distinguishing individuals with T1DM from those with T2DM in a setting with a large number of individuals with T1DM.

We estimated that nearly two-thirds of incident dementia among adults with T1DM could be attributed to their diabetes status. T1DM is not common, so it accounts for a fairly small fraction of all dementia cases, but for the growing number of individuals with T1DM over 65, these findings underscore the urgency of identifying and intervening on the mechanisms linking T1DM to dementia.^37,38^

Our findings regarding T2DM are consistent with prior literature. T2DM may contribute to dementia etiology through several mechanisms, including hyperglycemia, increased beta-amyloid deposition, or comorbidities such as metabolic syndrome, hyperinsulinemia, or stroke.^39–41^ In contrast, T1DM is characterized by pancreatic beta-cell destruction and the need for insulin supplementation to address hypoglycemia. Hypoglycemic events may increase dementia risk through neuronal damage caused by altered glucose metabolism and insulin insufficiency, or via oxidative stress and inflammation in the hippocampus.^42–45^ Whereas T2DM entails insulin resistance and hyperglycemia, T1DM may present a unique set of risks, in part due to the autoimmune destruction of beta cells. These mechanisms should be further explored.

With more than 5,000 participants identified with T1DM, our study is among the largest to examine the association between T1DM and dementia risk, although a prior study in England included over 340,000 individuals with T1DM. Our analyses were feasible because we developed and validated an algorithm to accurately distinguish individuals with T1DM from those with T2DM. Our diabetes classification integrates multiple sources of information including diagnostic codes, laboratory measures, and self-report information, which could help capture the complexities of diabetes presentations and treatments, improving classification accuracy.

Our study has some limitations. First, our assessment of all-cause dementia relied on EHR based diagnoses, which may entail misclassification. Dementia is often underdiagnosed and oftentimes delayed in routine clinical care. That said, individuals with T1DM may have more frequent contact with the healthcare system, which could increase their likelihood of earlier dementia diagnoses relative to those without DM. Second, while our algorithm for identifying T1DM demonstrated good sensitivity, even modest misclassification could bias our estimates given that T1DM is rare. For each reference standard, we selected a classification threshold that maximized Youden’s J index to balance sensitivity and specificity; however, this threshold may not be optimal for all analytic purposes and may depend on the research question at hand. Misclassification of DM status could have resulted from incomplete information. For example, AoU participants without diabetes-related EHR encounters were classified as having no diabetes, but participants’ EHR data may not capture care received from multiple networks. However, we believe that it was unlikely that individuals without DM were misclassified as having DM, particularly not T1DM. Finally, AoU’s recruitment process, largely convenience sampling through academic institutions and affiliated clinics, may limit generalizability. In a recent study comparing AoU with the nationally-representative NHANES, associations between various characteristics and all-cause mortality were in the same direction across the two studies, but some associations, especially those for clinical characteristics, were stronger in AoU.^46^

This study provides strong evidence that both T1DM and T2DM are associated with an elevated risk of dementia, underscoring the need to consider diabetes subtype when evaluating long-term cognitive outcomes. By developing and validating a classification algorithm to distinguish T1DM from T2DM using EHR data, we addressed a critical methodological need for more accurate surveillance of diabetes-related dementia risk in large population-based datasets. Our findings highlight the substantial burden of dementia among individuals with T1DM—particularly relevant as this population continues to age—and suggest the need for targeted prevention strategies.

Future research should explore the distinct biological mechanisms linking each diabetes subtype to dementia and quantify the impact of misclassification bias in studies relying on EHR-based diabetes phenotyping.

## Data Availability

The data and relevant code used in this study are made available to researchers who have access to the All of Us registered and controlled data tiers via the Research Hub.

