## Supplementary Material for "Type I diabetes and incident dementia: a prospective study in the All of Us cohort"

**eMethods**

**eFigure 1.** Process of developing the diabetes classification algorithm

**eTable 1.** Definition of Three Reference Standard Measures for Diabetes Type Classification and sample sizes of each used to validate the algorithm

**eTable 2.** Sensitivity and Specificity of Classifying Type 1 Diabetes Mellitus Across Five EHR Encounter Thresholds and Three Reference Standard Measures

**eTable 3.** Dementia hazard ratio specific to type of diabetes mellitus (reference: no diabetes mellitus), all participants and by gender, where T1DM was defined based on three or more EHR encounters

**eTable 4.** Population Attributable Fractions (PAFs) and Attributable Fractions (AFs)

**eTable 5.** List of Health Care Provider Organizations

**eTable 6.** Personal and Family History Survey Self-Report Diabetes Questions

**eTable 7.** SNOMED Codes – Dementia

**eTable 8.** SNOMED Codes – Type 1 Diabetes Mellitus

**eMethods**

***Algorithm for Classifying Diabetes Type***

*Personal and Family Health History Survey*

Under the ‘Hormone and Endocrine Conditions’ section of the Personal and Family Health History survey, participants were asked, *‘Have you or anyone in your family ever been diagnosed with the following hormone and endocrine conditions? Think only of the people you are related to by blood. Select all that apply.’* Within this section, there was an option for T1DM, where participants could check ‘Self’ as applicable. If a participant checked ‘Self’, they were then posed additional questions about their T1DM diagnosis, including questions about whether they were seeing a doctor or health care provider for T1DM, age of diagnosis for T1DM, and whether they were currently prescribed and/or taking or receiving treatment for T1DM. The same questions were asked to participants who selected ‘Self’ if they had been diagnosed with T2DM. Participants were able to endorse both T1DM and T2DM.

**eFigure 1. Process of developing the diabetes classification algorithm**

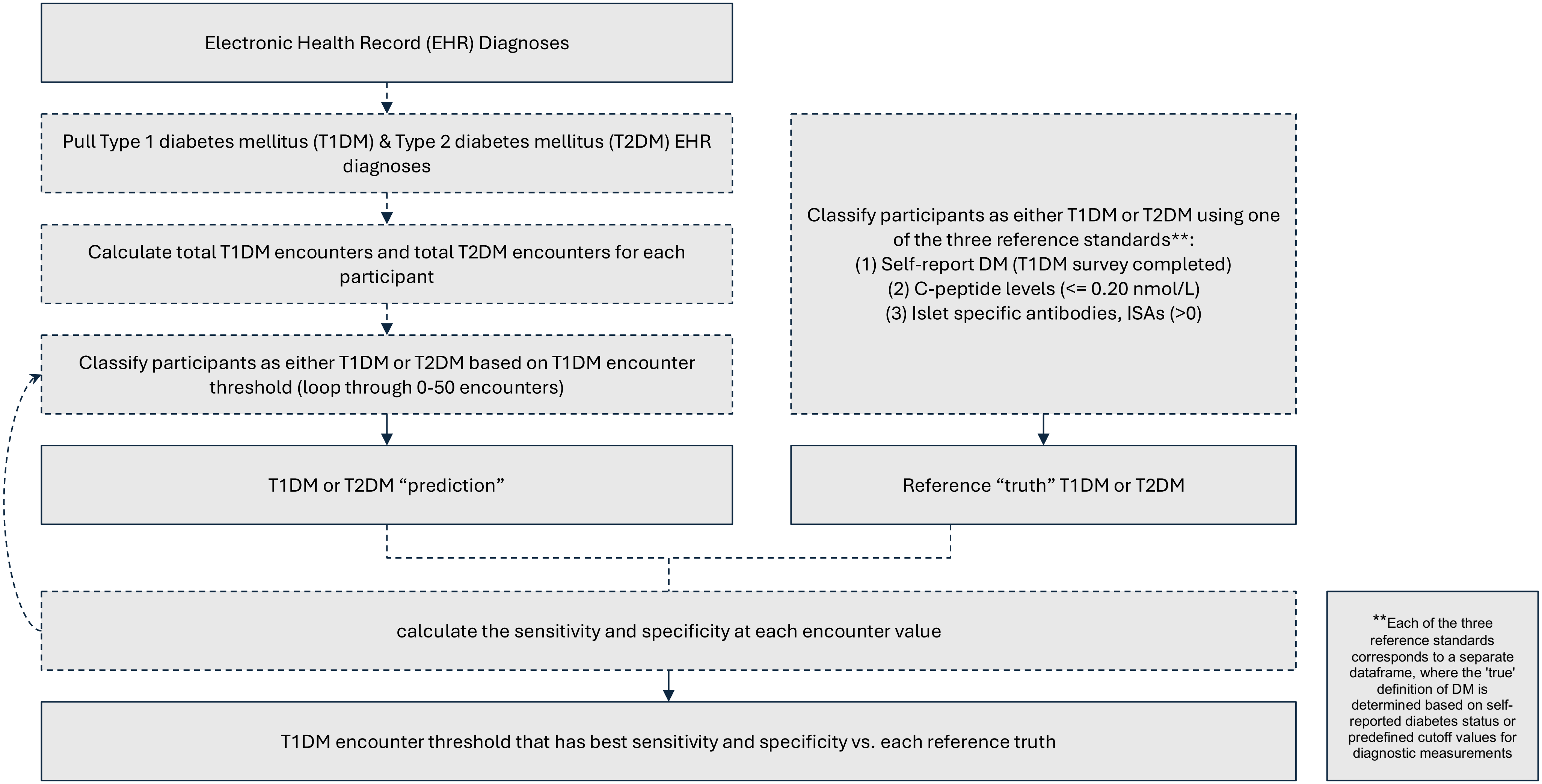

**eTable 1. Definition of Three Reference Standard Measures for Diabetes Type Classification and sample sizes of each used to validate the algorithm.**

| Gold Standard (Truth) | Diabetes Type | Definition | n |
|---|---|---|---|
| Self-Report Diabetes | Type 1 Diabetes | Completed the self-report type 1 survey (could have completed the type 2 survey) | 2,342 |
| | Type 2 Diabetes | Completed only the self-report type 2 survey | 17,376 |
| C-peptide Values | Type 1 Diabetes | Low C-peptide (<= 0.20 nmol/L) | 166 |
| | Type 2 Diabetes | C-peptide measurement other than low (between 0.20 and 0.30, or above 0.3) | 807 |
| Islet-autoantibody | Type 1 Diabetes | Had a positive value for an islet antibody | 310 |
| | Type 2 Diabetes | Had value of 0 for islet antibody | 293 |

**eTable 2. Sensitivity and Specificity of Classifying Type 1 Diabetes Mellitus Across Five EHR Encounter Thresholds and Three Reference Standard Measures**

| Reference Standard Measure | Threshold | Sensitivity | Specificity |
|---|---|---|---|
| Self-reported diabetes | 1 | 0.592 | 0.904 |
| | 2 | 0.527 | 0.950 |
| | 3 | 0.492 | 0.966 |
| | 4 | 0.464 | 0.975 |
| | 5 | 0.444 | 0.977 |
| C-peptide value | 1 | 0.837 | 0.674 |
| | 2 | 0.783 | 0.737 |
| | 3 | 0.765 | 0.791 |
| | 4 | 0.722 | 0.821 |
| | 5 | 0.710 | 0.836 |
| Islet-specific antibody | 1 | 0.583 | 0.567 |
| | 2 | 0.542 | 0.661 |
| | 3 | 0.505 | 0.700 |
| | 4 | 0.481 | 0.745 |
| | 5 | 0.470 | 0.751 |

Threshold refers to the number of T1DM labeled EHR encounters or more required for classification as T1DM.

**eTable 3. Dementia hazard ratio specific to type of diabetes mellitus (reference: no diabetes mellitus), all participants and by gender, where T1DM was defined based on three or more EHR encounters**

| | | Dementia hazard ratio (95% confidence interval), diabetes type (type 1 diabetes, type 2 diabetes, no diabetes [referent]) | |
|---|---|---|---|
| | Dementia cases /Total | Adjusted for demographic characteristics[a] | Additional adjusted[b] |
| No diabetes (referent) | 1452/2348 | 1.00 | 1.00 |
| Type 1 diabetes mellitus[c] | 68/2348 | 2.66 (1.98, 3.55) | 2.16 (1.51, 3.09) |
| Type 2 diabetes mellitus[c] | 828/2348 | 1.93 (1.73, 2.15) | 1.82 (1.60, 2.08) |
| **Gender** | | | |
| **Female** | | | |
| No diabetes (referent) | 769/1245 | 1.00 | 1.00 |
| Type 1 diabetes mellitus[c] | 40/1245 | 2.74 (1.84, 4.08) | 2.37 (1.46, 3.85) |
| Type 2 diabetes mellitus[c] | 436/1245 | 2.03 (1.74, 2.36) | 1.90 (1.57, 2.29) |
| **Male** | | | |
| No diabetes (referent) | 658/1060 | 1.00 | 1.00 |
| Type 1 diabetes mellitus[c] | 27/1060 | 2.54 (1.66, 3.90) | 1.93 (1.13, 3.29) |
| Type 2 diabetes mellitus[c] | 375/1060 | 1.84 (1.58, 2.15) | 1.77 (1.48, 2.12) |

a] Adjusted for sociodemographic factors: age, gender, race and ethnicity, educational attainment and household income. [b] Fully adjusted models were further adjusted for the following mid-life factors: smoking history, and alcohol use. [c] Reference group is no DM; type 1 diabetes includes individuals who have any form of T1DM (either T1DM alone or T1DM and T2DM); type 2 diabetes includes only individuals with T2DM alone.

**eTable 4. Fraction of dementia cases that would be eliminated if the dementia incidence of people with Type 1 Diabetes Mellitus (T1DM) matched that of individuals without diabetes, among sample members with T1DM (attributable fraction [AF]), and in the whole study population (population attributable fraction [PAF]), overall and by gender, and race and ethnicity.**

| | Prevalence of T1DM (%) | Prevalence of T1DM (%) among individuals with AD/ADRD | Dementia hazard ratio (T1DM vs no DM) | PAF (%) | AF (%) |
|---|---|---|---|---|---|
| Overall | 1.97 | 6.05 | 2.82 | 3.90 | 64.53 |
| Male | 0.98 | 5.66 | 2.59 | 3.47 | 61.38 |
| Female | 0.93 | 6.58 | 3.04 | 4.41 | 67.10 |
| White | 0.94 | 4.32 | 2.98 | 2.87 | 66.44 |
| Hispanic/Latino | 0.39 | 11.79 | 3.29 | 8.20 | 69.60 |
| Other | 0.65 | 7.05 | 2.38 | 4.08 | 57.98 |

Estimates are based on the prevalence of T1DM among participants who developed dementia in the All of Us sample used in this study and use hazard ratios from the sociodemographic adjusted model of T1DM and incident dementia. Population attributable fractions (PAFs) were calculated using Miettinen's formula.

**eTable 5. List of Health Care Provider Organizations**

| Health Care Provider Organizations (HPO) | Location |
|---|---|
| ***All of Us* Arizona** | |
| Banner Health | AZ |
| University of Arizona | AZ |
| **Trans-American Consortium for Health** | |
| Baylor Scott and White Health | TX |
| Corewell Health | MO |
| Essentia Health | MN |
| HealthPartners Institute | MN |
| Henry Ford Health | MI |
| Reliant Medical Group | MA |
| Saint Louis University | MO |
| University of Massachusetts Medical Center | MA |
| ***All of Us* New England** | |
| Boston Medical Center | MA |
| Massachusetts General Hospital | MA |
| ***All of Us* California** | |
| Cedars-Sinai Medical Center | CA |
| El Centro Regional Medical Center | CA |
| UC Davis Health | CA |
| UC San Diego Health | CA |
| UC San Francisco Health | CA |
| **Cherokee Health Systems** | TN |
| **Community Health Center, Inc.** | CT |
| ***All of Us* Southern Network** | |
| Cooper Green Mercy Health Services | AL |
| LSU Health Sciences Center New Orleans | LA |
| Tulane University | LA |

| | |
|---|---|
| UAB Medicine | AL |
| UAB School of Medicine, Montgomery Regional Medical Campus, Huntsville Regional Medical Campus, Selma Family Medicine Program | AL |
| University of Alabama University Medical Center | AL |
| University of Mississippi Medical Center | MS |
| University of South Alabama | AL |
| **Cooperative Health** | SC |
| ***All of Us* Puerto Rico** | |
| COSSMA | PR |
| Puerto Rico Consortium for Clinical Investigation | PR |
| University of Puerto Rico Comprehensive Cancer Center | PR |
| ***All of Us* Southern California Consortium** | |
| Memorial Care | CA |
| UC Irvine Health | CA |
| University of Southern California | CA |
| **Illinois Precision Medicine Consortium** | |
| Endeavor Health | IL |
| Northwestern Medicine | IL |
| Rush University Medical Center | IL |
| The University of Chicago | IL |
| University of Illinois at Chicago | IL |
| ***All of Us* Wisconsin** | |
| Froedtert & Medical College of Wisconsin | WI |
| Gundersen Health System | WI |
| Marshfield Clinic Health System | WI |
| University of Wisconsin School of Medicine and Public Health | WI |
| Loma Linda University Health | WI |
| ***All of Us* Southeast Enrollment Center** | |
| Morehouse School of Medicine | GA |
| **Mount Sinai Health System** | NY |

| | |
|---|---|
| **The University of Miami Miller School of Medicine** | FL |
| ***All of Us* New York City** | |
| New York-Presbyterian Hospital | NY |
| NYC Health + Hospitals/Harlem | NY |
| Weill Cornell Medicine | NY |
| **San Ysidro Health** | CA |
| **Sun River Health** | NY |
| ***All of Us* Heartland** | |
| The University of Kansas Health System | KS |
| University of Iowa Health Care | IA |
| University of Kansas Medical Center | KS |
| University of Missouri | MO |
| University of Nebraska Medical Center | NE |
| **UT Health Science Center at Tyler** | TX |
| **U.S. Department of Veterans Affairs** | CA/MA |
| **University of California, Los Angeles** | CA |
| ***All of Us* North Carolina** | |
| University of North Carolina, Chapel Hill | NC |
| University of North Carolina, Nutrition Research Institute | NC |
| ***All of Us* Pennsylvania** | |
| University of Pittsburgh | PA |
| **VCU Wright Center for Clinical and Translational Research** | VA |
| **Waianae Coast Comprehensive** | HI |
| **Yale University** | CT |

**eTable 6. Personal and Family History Survey Self-Report Diabetes Questions**

**Hormone and Endocrine Conditions**

Have you or anyone in your family ever been diagnosed with the following hormone and endocrine conditions? Think only of the people you are related to by blood. Select all that apply.

**Type 1 diabetes**

Including yourself, who in your family has had Type 1 diabetes? Select all that apply.

Self

*Branching logic: when "Self" selected, then:*

Are you still seeing a doctor or health care provider for Type 1 diabetes?

About how old were you when you were first told you had Type 1 diabetes?

Are you currently prescribed medications and/or receiving treatment for Type 1 diabetes?

**Type 2 diabetes**

Including yourself, who in your family has had Type 2 diabetes? Select all that apply.

Self

*Branching logic: when "Self" selected, then:*

Are you still seeing a doctor or health care provider for Type 2 diabetes?

About how old were you when you were first told you had Type 2 diabetes?

Are you currently prescribed medications and/or receiving treatment for Type 2 diabetes?

**eTable 7. SNOMED Codes – Dementia**

| Code | Definition |
|---|---|
| 191519005 | Dementia associated with another disease |
| 26929004 | Alzheimer's disease |
| 1591000119103 | Dementia with behavioral disturbance |
| 15662003 | Senile dementia |
| 12348006 | Presenile dementia |
| 191493005 | Drug-induced dementia |
| 278857002 | Dementia of frontal lobe type |
| 428051000124108 | Mild dementia |
| 429998004 | Vascular dementia |
| | Subcortical dementia |

**eTable 8. SNOMED Codes – Type 1 Diabetes Mellitus**

| Code | Definition |
|---|---|
| 46635009 | Type 1 diabetes mellitus |
| 313435000 | Type 1 diabetes mellitus without complication |
| 199229001 | Pre-existing type 1 diabetes mellitus |
| 609566000 | Pregnancy and type 1 diabetes mellitus |
| 190368000 | Type 1 diabetes mellitus with ulcer |
| 426875007 | Latent autoimmune diabetes mellitus in adult |
| 31321000119102 | Diabetes mellitus type 1 without retinopathy |
| 23045005 | Insulin dependent diabetes mellitus type 1A |
| 28032008 | Insulin dependent diabetes mellitus type 1B |
